# Sickle Cell Disease Screening: Experiences of Parents with Positive Newborns at Korle Bu Teaching Hospital

**DOI:** 10.1101/2025.11.11.25338668

**Authors:** Irene Kanyoke, Faustina Hayford Blankson, Diana Dwuma-Badu, Catherine Segbefia

## Abstract

**Objective.:** To explore parents’ experiences following receipt of positive newborn screening for sickle cell disease (SCD) test results at a teaching hospital in Ghana.

**Methods.:** Using purposive sampling, data were collected using semi-structured individual guides to interview parents of newborns attending the paediatric SCD clinic at Korle Bu Teaching Hospital (KBTH), the major referral health facility in Ghana. Questions were categorized into 3 main areas: parents’ reactions to initial positive results, parents’ responses to in-clinic counselling, and parents’ attitudes towards follow-up clinic care. Interviews were conducted face to face either in English or a local dialect (Twi) after which they were transcribed verbatim. Data were analyzed using NVivo software to identify relevant codes which were organized into themes and sub-themes.

**Results.:** Twenty-four mothers and one father were interviewed between 2nd November 2021 and 28th January 2022. The ages of their children ranged from 7 months to 3 years 9 months. Seventeen (68%) of the parents were married, 5 (20%) had more than one child enrolled in the clinic, and 10 (40%) reported that they or their partners had SCD. The 5 themes identified were: (i) emotional response after disclosure, (ii) decision to share results, (iii) care and management of newborns at home, (iv) knowledge of genetics of SCD, and (v) adherence to routine follow-up. Positive test results elicited worry, surprise, or shock in the majority, especially those with no previous knowledge of or experience with SCD. Most parents had received counselling and educational information on caring for their positive newborns at home and the danger signs of SCD. The decision to share test results with others was influenced by fear of stigmatization. While desiring to see their babies remain healthy, parents defaulted on clinic visits due to financial difficulties and the COVID-19 pandemic.

**Conclusion.:** Knowledge of SCD and personal experience influenced parents’ reaction to initial positive newborn screening for SCD test results. There is a need for more education about SCD among the general population and institutions of strategies to provide psychological and socioeconomic support for families attending the KBTH clinic.

## Introduction

Sickle Cell Disease (SCD) is a hereditary disorder characterized by the presence of abnormal haemoglobin the most common of which is the haemoglobin S (Hb S) [1]. SCD clinically presents as acute haemolytic and vaso-occlusive crises which can lead to complications including stroke, organ damage, priapism and acute chest syndrome, among others and it is estimated that between 300,000 to 500, 000 babies are born with SCD worldwide in a year [2]. Studies show that 7 in 10 SCD-related deaths are preventable and the high prevalence of morbidity and mortality in Africa is made worse by inadequate systemic screening for the disease, inadequate follow-up care, shortage of facilities for SCD management, poor research and intervention protocols [3,4].

SCD has a huge socio-economic impact not only on parents but also on the community and even the country at large [3] and as such, over the years, there have been several disease management interventions for the disease. One of these interventions is Newborn Screening (NBS). Newborn screening for SCD is the process by which blood from a baby is collected in the first few days after birth and tested for SCD [5]. The main objective of newborn screening is to allow for the early identification of infants with the disease and the provision of follow-up care to help reduce morbidity and mortality [6]. Newborn screening for SCD is a laudable idea and comprehensive follow-up care which involves a multi-disciplinary approach must include the families and caregivers of newborns with SCD [5].

In Ghana, universal newborn screening however is not being fully implemented [7] and in the few places where screening is taking place, barriers to care include parents refusing to accept an initial positive diagnosis or failing to attend the clinic after the initial visit [8]. According to unpublished records from the Newborn Screening programme in KBTH, a total of 370 newborns with P-SCD were identified through screening out of a total of 20,167 newborns screened at KBTH and Ussher Hospital from June 2017 to December 2022 representing 1.8% of the total screened [9,10]. Even though a total of 336 of these P-SCD babies were tracked and parents were given the results, 239 parents showed up for the initial visit with an average enrolment age of 6 months while 78 parents (28% of those contacted) refused to accept the initial diagnosis [9,10]. There were also challenges with mothers failing to attend clinic after the initial visit, only coming back after their child had become ill/symptomatic [9]. This defeats the main principle of newborn screening where the aim is to start penicillin prophylaxis and comprehensive follow-up care by 3 months of age.

Due to these aforementioned deficiencies, this study aimed to explore the experiences of parents of newborns following the receipt of positive NBS for SCD results at a teaching hospital in Ghana.

This study used an adaptation of Urie Bronfenbrenner’s Social-Ecological Model (SEM) developed in 1979. The SEM has been adopted by public health experts over the years for research into understanding issues affecting well-being. Thus, this framework was adapted for this study to better explore the multifaceted factors that affect parents’ experiences. See Figure 1 for an illustration of the modified Social-Ecological Model of Health. This study used an adapted version that examined the interplay and interconnectedness of individual (intrapersonal) and interpersonal factors that affect the experiences of parents of positive SCD newborns at the Paediatric Sickle Cell Clinic of Korle Bu Teaching Hospital. This model was used to study how the factors at each level interact and may influence parents’ experiences.

The main objective of this study was to explore the experiences of parents of positive sickle cell disease newborns at the Paediatric Sickle Cell Clinic (PSCC) of Korle Bu Teaching Hospital.

The specific objectives were:

1. To explore the reactions of parents of newborns towards disclosure of positive SCD results at the PSCC of KBTH.
2. To explore the responses of parents of P-SCD newborns to in-clinic counselling and support at the PSCC of KBTH.
3. To describe the attitudes of parents of SCD-positive newborns towards subsequent follow-up clinic care at the PSCC of KBTH.

## Method

This study used a qualitative research design with an explorative approach with data collection done using a semi-structured interview guide to conduct individual in-depth interviews. The study was conducted at the Paediatric Sickle Cell Clinic of the Korle Bu Teaching Hospital (KBTH) and the study population consisted of mothers and or fathers of newborns enrolled in the Pediatric Sickle Cell Clinic of KBTH. Inclusion criteria were all mothers or fathers of newborns diagnosed via the KBTH NBS programme and enrolled in the clinic while the exclusion criteria were Parents of newborns between 6 weeks to 4 years old, diagnosed via the KBTH NBS programme and enrolled in the clinic but attending the clinic for the first time. Parents of newborns, diagnosed via the KBTH programme, enrolled in the clinic but less than 6 weeks or more than 4 years were also excluded.

The researchers used the purposive sampling method to recruit participants. According to Francis et al, data saturation for theory-based qualitative studies can be decided by using 2 principles: 1. establishing an initial minimum sample size for analysis and 2, specifying the stopping criterion i.e., how many more interviews will be conducted without new ideas [11]. Before starting data collection, a total of thirty-six (36) interviews were expected to be conducted, continuing until saturation. However, due to time constraints and restrictions due to the COVID-19 pandemic, the sample size was modified to eighteen (18) with the stopping criterion being three (3). Saturation was achieved after twenty-four (24) parents were interviewed. The researcher used 2 interview guides: a questionnaire to collect socio-demographic information and a semi-structured individual interview guide. The interview guide was piloted at the Paediatric Sickle Cell Clinic of KBTH once approval had been received from the Institutional Review Board of Korle Bu Teaching Hospital after which modifications were made. The semi-structured individual interview guide was developed using the conceptual framework and objectives of the study and included open-ended questions and socio-demographic questions. These interviews were conducted at the PSCC venue during clinic days and each interview took between 40 minutes to an hour and was conducted in Twi or English and tape-recorded for transcription later. Thirteen interviews were conducted in English while eleven were conducted in Twi. NVivo software was used to assist in the analysis and coding of data. To ensure the privacy and safety of the data collected, all audiotape recordings, field notes and memos were kept under lock and key and only accessible to authorized persons.

Sufficient controls were put in place to preserve the safety of the data as well as to guarantee its accuracy devoid of any bias. As such, the researchers used various validation strategies to ensure credibility and trustworthiness. Credibility was achieved in this study by using a codebook, which served as a trail of evidence for all data gathered and analysed. A field diary and a memo were also used to record all observations during interviews and analysis of data. Data were triangulated using memo notes, field diaries and transcriptions from interviews in the write-up to ensure trustworthiness and credibility. Saturation was achieved in this study by utilizing the 2 principles of achieving saturation according to Francis et al, [11] where the initial minimum sample size and stopping criterion were established before data collection.

Another way of ensuring trustworthiness and credibility in this study was the use of 2 peer-debriefers in the person of my academic supervisor Dr Faustina Blankson and Ms Diana Dwuma-Badu, a Haematologist nurse specialist, and a researcher at the sickle cell clinic of KBTH. Transferability, another quality of validity was achieved by using rich, thick descriptions and quoting participants’ voices under each theme.

The researcher used an iterative process of checking interpretations and analysis of data with the peer-debriefer as well as constantly comparing the responses of participants to help influence the validity of the study.

Data Analysis using thematic analysis was done after data collection had been concluded. The transcripts from the pilot were not added to those from the main study. Audiotaped interviews from the main study were transcribed verbatim after each interview and those conducted in the local languages were then translated into English. Thematic Analysis following the 6 phases of thematic analysis as defined by Braun and Clark was used to establish emerging themes.

Open coding (codebook) using an inductive approach was used as a first step in identifying initial codes. Similar codes were then clustered to form categories after which the relationships between the emerged codes were analysed and categorized into themes. Ethical approval for this study was sought and acquired from the Institutional Review Board of Korle Bu Teaching Hospital (KBTH).

Consent was sought from participants before interviews began. Consent forms were given to parents to read and sign, while those who could not read had the consent translated verbally into the local language for them, after which they thumb-printed their consent. Participants’ anonymity was ensured by assigning pseudonyms to them and assuring them their real names would not be used.

All information gathered was treated with the utmost confidentiality. The recordings and other research materials will be appropriately discarded after 5 years.

## Results

The themes have been presented using constructs from the Socio-Ecological Model of Health. A total of twenty-four (24) mothers and a father of newborns who had tested positive for SCD at birth and enrolled in the clinic were interviewed. Four of the participants had babies who were less than one year old. Four more participants had children aged one year to two years and sixteen had children aged two years one month to three years, and eleven months.

The participants consisted of seventeen (17) married and three unmarried parents. Two other participants were not married but were in a relationship with the father of their babies while two were separated from their husbands. All participants lived within the Greater Accra Region. Nine of the participants had completed tertiary education, three had completed primary six, five had completed Junior High School (JHS), six had completed vocational or Senior High School (SHS), and one had not had any formal education at all. Five of the participants had more than one child enrolled in the clinic. Ten (10) of the participants also had the disease or had partners who had the disease while fourteen (14) participants had the SCD trait. More than half of the participants reported earning less than 500 cedis for the household per month. This reflects the socio-economic costs of the disease and provides insight into how socio-economic challenges influence parents’ experience at the clinic. The results are found in Table 1.

The emerged codes were categorised using the socio-ecological model indicating the experiences of parents of positive newborns. The codes were categorised into 2 main constructs based on the socio-ecological framework, namely: intrapersonal influences and interpersonal influences. A summary of the codes and themes can be found in the codebook in Table 2. The results are presented as follows: Intrapersonal influences (Emotional Response after Disclosure, Decision to Share Results, and Adherence to Routine Clinic) and Interpersonal influences (Care and Management of the Disease at Home, knowledge of Genetics of SCD). In total, there were 5 main themes and 21 sub-themes.

### 1.0 Intrapersonal Influences

Three themes emerged from this construct: Emotional response after disclosure, the decision to share results and adherence to routine clinic. Emotional response after disclosure had 5 sub-themes namely: scared, worried, not surprised, disbelief and surprise/shock. Under the decision to share results, there were 3 sub-themes: shared results with partner only, fear of social exclusion stigma and shared results with family and friends. The third theme under intrapersonal influences was adherence to routine clinic and under this theme, there were 6 sub-themes: adherence to routine visits, adherence due to parents’ personal experience with the disease, non-adherence due to COVID-19, non-adherence due to financial difficulties, adherence to daily medication and routine lab tests.

### 1.1 Emotional Response After Disclosure

#### Scared

Participants reported that they felt scared when they were initially told of the positive results. For participants, this reaction was due to the little knowledge or preconceived ideas they had about the disease.

> *“You know, life is hard now, and I was scared that if it happens that she is a sickler, what would I do?”*
>
> *(Female Participant, in her 40s)*
>
> *“It’s not easy oo. It’s not easy ((with tears in voice)). Me knowing how it is. Especially, me, if I didn’t even know how it is. But me knowing how the pain, hmmm, the, the, hmm, it’s not easy oo. I’m facing a lot inside me… And I’ve been keeping this in me. And I don’t know what to do. I don’t know what to do. Hmm, it’s really, difficult thing, ever since I got to know my son, has this disease.”*
>
> *(Female participant, in her 20s)*

#### Worried

Some participants also reported being worried after receiving the results. They reported they got worried because of the knowledge that sickle cell disease had a lot of socio-economic ramifications. As such, a positive diagnosis was very worrying for them.

> *“Because I can sometimes look at the child, and shake my head and think to myself “This disease too where is it coming from”? That I gave birth and need to be at the hospital every 3 months. It’s a lot of worry.”*
>
> *(Female participant, in her 30s)*

##### Not surprised

Some of the participants on the other hand had not been surprised by the positive results of their newborns as they already had prior knowledge of the family history of the disease and as such, knew of the possibility of their baby testing positive for the disease. Thus, they were not surprised by the result.

> *“I wasn’t surprised when they told me. Knowing that my other girl already had the disease. I understood.”*
>
> *(Female participant, in her 30s)*
>
> *“So him ((pointing to child with SCD at clinic)), when his results came, I wasn’t surprised because I knew the dad is SC. so I wasn’t surprised at all.”*
>
> *(Female Participant, in her 30s)*
>
> *“I wasn’t really surprised. Because I had been told that if I give birth to more children, all my children could have it.”*
>
> *(Female participant, in her 40s)*

##### Disbelief

Some participants also, reported that they did not believe the results when they were initially told about the results. This was because they were not expecting the results. Some participants, however, then came to believe and accept the positive results when their children started showing symptoms of the disease.

> *“And then later she called and informed us that he had the disease, but I didn’t believe it. And then a few months later, he started getting sick.”*
>
> *(Female participant, in her 30s)*
>
> *“That first day they gave me the results, I didn’t believe it.”*
>
> *(Female participant, in her 30s)*

However, some of the participants reported that even after receiving the positive results and having attended the clinic for a while they and or their partners still did not believe the results.

> *“no. he still doesn’t believe. He said he donates blood to people all the time. So if he had the disease, they wouldn’t allow him to.”*
>
> *(Female Participant, in her 30s)*
>
> *“So it’s only my husband and I that know. And we don’t believe in that thing, so, we don’t see our child as SC.”*
>
> *(Female participant, in her 20s)*

##### Shocked/surprised

There were however several participants who reported being shocked or surprised by the results even though they had prior knowledge of the disease and knew that they had the trait and could therefore give birth to a child with the disease.

> *“It did help. Because even before we got married, I was contemplating. And then when the news came, even though it came as a shock, I knew, this was what was before us. And then. So, it was quite okay.”*
>
> *(Female participant, in her 30s)*

### 1.2 Decision to Share Results

#### Shared results with a partner only

Some, however, reported that apart from their partner or the child’s father, they had not shared the results with any other person. They stated they had kept the results just between the 2 of them and not shared it with any other person.

> *“Well, I didn’t want anyone to know. I just wanted it to be between me and my husband, so I didn’t tell anyone.”*
>
> *(Female participant, in her 40s)*

#### Fear of social exclusion and stigma

Some of the reasons parents gave for not sharing the results with extended family and friends included fear of judgment, social exclusion and stigma.

> *“Hmm, for my family, I don’t really mind telling them, but it’s because I don’t want people to say the child is this or the child is that. Yes, I don’t want someone to stand somewhere and tell another person about it and say something like oh this child is beautiful but ahh, she has this disease. And then it will reduce her marks or something. I don’t want that. Yes, you see people will talk. Maybe they don’t mean anything bad but,”*
>
> *(Female participant, in her 40s)*
>
> *“Nobody oo. I’ve not told anybody. Because of the stigma, I don’t want to, to tell anybody… Because of what I went through, I don’t want to tell people about it. My mom didn’t keep quiet, and people would tease me and all that. So, I don’t want that.”*
>
> *(Female participant, in her 30s)*

#### Shared results with family and friends

Some parents, however, did not mind sharing the results with some family and friends as they felt those people were closer to them and were part of providing care for the children. As such, they felt they needed to know to help them care for the child.

> *“Yes. I told the caretaker. Because if I have to leave, and you don’t give the medication, I will blame her. And then recently, I told my mother-in-law… and then my parents. My siblings are also aware. So that in case there is something to give, I tell you, chilled things are not for her. I’ll tell you please, hold on.”*
>
> *(Female participant, in her 30s)*

### 1.3 Adherence to Routine Follow-up Procedures

#### Adherence due to the expectation for a healthy outcome for the child

Parents reported attending the clinic regularly because they wanted their children to remain healthy.

> *“I’ve been coming to the clinic for the past 7 to 8 months. Every month. I’m here. Yes, sometimes even 2 weeks. In a month, I can come like 2 times … for me, I always say that because of his health. I want him to be healthy. And for his breathing to be fine. Because sometimes, when we come, they check everything, his breathing, and they do a physical exam of him to make sure everything is fine. So, I bring him to make sure everything is fine. Because when we are home, everything is fine. But, when I’m given a date to come, I have to come.”*
>
> *(Female Participant, in her 30s)*

#### Adherence due to parents’ personal experience with the disease

Others also reported adhering to regular monthly or bi-monthly clinic visitation due to previous experience of parents living with the disease and as such found it necessary to keep the child healthy.

> *“Because I have that experience. So I don’t want her to pass through the stress I have gone through, so I come to the clinic.”*
>
> *(Female participant, in her 30s)*

#### Non-adherence due to financial difficulties

There were, however, some who reported that they sometimes missed appointments due to circumstances such as financial difficulty.

> *“Madam please, you see, for my son, I love him. And it’s the doctor who knows, so whatever they tell me I have to listen and do it. But sometimes, when I’m unable to come on the date, it’s usually because of financial difficulties. I may be lacking money for the labs …. But financially, it’s been difficult. Because of money issues. I was supposed to bring him last month but because I didn’t have money, I couldn’t come.”*
>
> *(Female participant, in her 40s)*

#### Non-adherence due to the COVID-19 pandemic

Some of them also reported that they did not attend the clinic for a while due to the COVID-19 pandemic. They reported that even though they were scheduled for regular clinic visitations, some of these circumstances prevented them from visiting the clinic regularly.

> *“We come. Every 3 months, we come. But we were asked to come and do some labs. But we couldn’t do it. Because of the COVID. I was scared so I didn’t come. And then I was called in November to bring her. And we will come again in February.”*
>
> *(Female participant, in her 30s)*

#### Adherence to daily medication

For the basic medication, all parents reported that they strictly followed the guidelines of daily medication for their children. Parents reported always refilling the medication and stated that they always made sure to give the child medication every day, as prescribed by specialists.

> *“Every day, I give her the medication. I follow all the instructions I’m given. I give her every day. So she is fine and she is very fast too.”*
>
> *(Female participant, in her 30s)*
>
> *“Every day. I give her the medication. Every day. When it finishes, I go back and buy. Yes, the other time I came, the doctor wrote another prescription with the same medication for me so, now I know it is the same medication I have to keep giving her. So once it finishes, I buy more. Once it runs out, I call my husband and if he has money, he sends it.”*
>
> *(Female participant, in her 40s)*

All parents also reported making sure to refill the prescription every time it finished.

> *“And so because of that, I don’t play with the medication. Once it finishes, I make sure I refill it. And he is fine. He has not been sick since. Once in a while he gets a cold, and when that happens, I take him to Amasaman, then they give us medication and we go back home. That’s it.”*
>
> *(Female participant, in her 30s)*

#### Routine Laboratory Tests

Parents reported that they were routinely asked to do lab tests such as full blood count, reticulocyte count, and sometimes liver function tests, among others. This they stated, was done routinely, most often once a month. Participants reported that they usually adhered to whatever lab requests they were given to do.

> *“And if I have to do any labs, if I have money, I hurry and do it, so the doctors can check his blood levels. So, I believe what they say is good for me.”*
>
> *(Female participant, in her 40s)*
>
> *“Oh it’s usually the lab we frequently do.”*
>
> *(Female participant, in her 40s)*

### 2.0 Interpersonal Influences

Themes under this construct emerged after looking at the network /relationship between parents and clinic staff and the counselling given to them. They reported that they had been educated on the care and management of their newborns at home as well as the genetics of SCD during counselling and educational sessions as part of comprehensive follow-up care provided at the clinic. Two themes emerged from this: Care and management of newborns with SCD and knowledge of the genetics of SCD.

Under care and management of newborns with SCD, 4 sub-themes emerged namely: preventive measures/care at home (nutrition, hydration, warm clothes), awareness of treatment options, awareness of possible complications and danger signs. There were 3 sub-themes which emerged under knowledge of the genetics of SCD and they were: inheritance of SCD, blood disorder and family history of the disease.

### 2.1 Care and Management of Newborns with SCD

#### Preventive Measures

Parents reported they were counselled on how to care for their newborns at home and how to take preventive measures to reduce illnesses and painful episodes at home.

They reported that they were counselled on how the nutrition or diet of the child is important and could improve the health of the child.

> *“They said to pay attention and take care of him well, his eating habits. The food he eats… They said I should pay attention to that. So that he doesn’t become sick. Yes.”*
>
> *(Female participant, in her 30s)*

Some parents also stated that they were advised to make sure the child drank lots of water and was always hydrated. Thus, they were advised on the importance of hydration for their children with SCD.

> *“Yes. I give him lots of fruits. He drinks water too. A lot. I make sure he wears pullovers and stuff.”*
>
> *(Female participant, in her 30s)*

Parents reported that they were also educated on the need to keep their babies warm.

> *“Hmm, they said many things. How to how to, eh, is it, prevent it, ehehn, in terms of not to take something cold. And all those kinds of things. You will know the way when it’s cold, you should put something, like a cardigan, all those kinds of things. And you have to take your medication always, no crises, for 0 months to 5 years. You won’t have any problem.”*
>
> *(Female participant, in her 30s)*

Medication literacy was one of the things imparted to parents as well and as such, parents reported that they were now aware of the medication as well as other treatment options necessary for managing the disease. Parents reported being advised to give their children penicillin, zinc supplements, folate supplements and hydroxyurea.

> *“He’s taking, umm, brufen, zincovit, folic acid, penicillin V. but recently he was given the*
>
> *… I’ve forgotten the name… yes. Hydroxyurea”*
>
> *(Male and female participants, both in their 20s)*
>
> *“And the doctor said not to think too much of it. So, when he keeps taking the medicine, small small it will become fine. Maybe the SS too can change. Because it’s good to treat them when they are little. Before they grow up.”*
>
> *(Female participant, in her 30s)*

#### Awareness of possible complications from the disease

Parents stated that they were counselled on the possible complications that could emerge from the disease. They reported that they were taught about how the disease could affect other parts of the body such as the eyes, the kidneys, etc.

> *“They told me the disease can affect other parts of the body. That’s why I was so sad when I couldn’t get the eye test for my son when his eye got swollen.”*
>
> *(Female participant, in her 40s)*

#### Awareness of danger signs

Parents informed us they were counselled on what danger signs to watch out for and actions to take to mitigate any symptoms at home. Parents were counselled and gained knowledge on watching out for danger signs such as yellow eyes, fever, swollen feet and hands, constant crying, etc.

> *“They said for sickle cell, if I see he is in pain or he gets a fever if his urine become coke coloured, I should bring him.”*
>
> *(Female participant, in her 20s)*
>
> *“They said he may get complications from the disease. When they are babies, maybe you will see one of their hands swelling or their feet swollen. Or their eyes. And when they get a temperature, you have to bring them to the hospital and not stay in the house. It might be an infection.”*
>
> *(Female participant, in her 30s)*

### 2.2 Knowledge of the Genetics of SCD

#### Inheritance of SCD

They reported that they were taught that the disease was inherited from both parents. Parents were made aware of the fact that they both had to have the trait to be able to have a child with the disease and that the disease was found in their genes and was passed down to their child. They also reported being informed of the possibility of having other children with the disease.

> *“Yes. The day I brought the results, the doctor asked that both my husband and I come. So, we came and he explained everything to us. They explained everything. That because my husband is AS and I’m AC, that’s how come. He took my husband’s S and took my C. The older one took my husband’s S and my A.”*
>
> *(Female participant, in her 30s)*
>
> *“They told us. Ehh. They told us that it’s genetic. If one of the parents has, then probably 1 or 2 of your children will have it.”*
>
> *(Female participant, in her 30s)*

#### Blood disorder

The parents also reported that they were educated on the fact that sickle cell disease is a blood disorder and that it is sickle cell shaped instead of round and breaks easily thus causing painful episodes.

> *“They took me through the sickling. How it forms. How their blood cells look like. Is it the white one or the, the red one? Yea, that looks like a bofloat, and theirs is like an arch, the moon, half-moon, which is very hard not like the others that are soft that can pass through all the blood. Theirs because is hard, sometimes, it gets broken and chokes the veins that you have to give them the (massage) and all that. So, I was taught all that.”*
>
> *(Female participant, in her 30s)*
>
> *“They told me that there is S in the child’s blood. And she showed me how it is, with a diagram, how the S looks in the blood, she drew it and showed me. And how that for us, the blood is not round but is curved. And that is how come it happens that way. And so, they will try and treat it. She said it is something that’s happening in Africa. Sickle cell disease. And she said so nowadays when they see that a child has the disease, they advise parents to bring the child to the hospital for care so that it can prevent certain complications.”*
>
> *(Female participant, in her 40s)*

#### Family history of the disease

Participants also stated that they were asked if they were aware of any known family member with the disease. They reported that they were informed that sickle cell was a familial disease passed down from parents to the child and could affect other family members.

> *“Because my father, for instance, he also has it … we are 3. My sister also has the disease. Hers is worse.”*
>
> *(Female participant, in her 30s)*
>
> *“It’s me and our last born (meaning youngest sibling) who have the disease in our family.”*
>
> *(Female participant, in her 40s)*

## DISCUSSION

### Objective 1 - Reactions of Parents to Disclosure of Initial Results

The first objective of this study was to explore how parents reacted when the initial positive results were disclosed to them. After data analysis, two themes emerged that provide an answer to this question: emotional response to the disclosure of results and the decision to share results.

Parents reported feeling scared, worried, disbelief and surprised or shocked due to all the negative information they had heard about the disease. As such they felt scared and worried that their children could also suffer from the disease. This doesn’t differ much from a study by Chudleigh et al [12] where they explored the experiences of parents of newborns receiving positive SCD carrier results. In that study, parents reported a range of emotions such as devastation, guilt, denial, surprise and shock. As much as these emotions may not have been the same, they were still negative emotions that were felt by parents receiving positive SCD results about their newborns.

Prior knowledge of SCD, especially the negative views could lead to doubt when parents are given a positive diagnosis [13]. In this study, parents’ reasons for disbelief were due to misconceptions and misinformation about the disease and as such, they believed that their children could not have the disease since their newborns looked very healthy.

One important point this study revealed was that prior knowledge of the disease and or personal experience with the disease has a huge impact on how parents react to positive results of SCD. Parents who had prior knowledge due to a family history of the disease stated that they were not surprised by the results while those who may have heard about the disease but did not know their carrier status were scared, worried, and surprised by the results of their newborns. Thus, prior knowledge of the disease impacts parents’ emotional reactions to the disclosure of positive SCD results of their newborns.

The decision to share the initial positive results was something that many parents did not take lightly due to issues such as stigma. In this study, most parents of newborns decided to share the positive results/ or diagnosis with someone, at least a partner with some preferring to keep the diagnosis just between them and their partners. Some parents’ decision not to share the results was based on the desire to not subject their child to stigma and also to prevent their child from being less socially accepted among friends and even family. A few of the parents however made decisions to share the results with their partners and a very few close family members.

This is in contrast with the study by Ulph et al [14] documenting parents’ responses to receiving sickle cell carrier results for their newborns, where parents felt a responsibility towards sharing the results with their extended family and parents had no struggle with informing extended families about the results. For others, the decision to share with at least just their partners was because of the expected support they could receive from them.

A study by Chudleigh et al [12] also contrasts the findings in this study as that study reported that most of the parents of newborns who had received positive SCD carrier results of their newborns shared results with family and friends. In this same study, however, some of the parents who had lived in Africa or had family back in Africa hesitated to inform that side of their family in Africa because they felt their families had a bleak outlook on the disease. It can therefore be said that geographical context and culture influence peoples’ experience of stigma and this in turn may influence parents’ decision to inform other people about the results of the newborns.

Social acceptance and stigma were thus a major part of the decision of parents to either share or not share the positive SCD results of their newborns.

### Objective 2-In-Clinic Counselling and Comprehensive Follow-up Care

The second research question aimed to look at how parents responded to in-clinic counselling and education and how this formed an essential part of comprehensive follow-up care. It aimed to provide insight into parents’ relationships and interactions with staff at the clinic in terms of the counselling and education provided by trained staff at the clinic. Analysis of data yielded two themes that answered this question: care and management of newborns with SCD and knowledge of the genetics of SCD.

Counselling patients with sickle cell disease is an essential tool in ensuring effective adjustment [15]. Educating parents on how to care for their children at home is one of the ways of reducing bacterial infections in children [16] Parents should be advised to give their children enough fluids to reduce dehydration. They should also be advised on wearing warm clothes in cold weather, and the need to ensure proper diet and nutrition [16,17].

In this study, parents of newborns were counselled and received a lot of knowledge on the nature of sickle cell disease, preventive measures and caring for their newborns at home. Parents were advised during counselling sessions with staff to make sure to take care of their newborns at home by ensuring they stayed hydrated, warm and had adequate nutrition as well as watching for danger signs such as yellow eyes, coke-coloured urine, bloated stomach, swollen joints, and excessive crying. Parents were also advised on the treatment options available for their children. As such, parents knew what medication they were prescribed and how that medication played a role in managing the disease. This is consistent with a study by Colah et al [18] documenting the Indian experience of newborn screening for SCD where parents were given daily folic acid and penicillin prophylaxis for their newborns. It is also consistent with the Jamaican experience study by King et al [19] where parents of newborns diagnosed via newborn screening were counselled on the need for routine penicillin prophylaxis. Medications such as folic acid, vitamin C, and penicillin have been known to reduce infections among infants with SCD.

Once a sickle cell diagnosis has been confirmed after the newborn screening, parents need to be educated on the type of SCD their child has, the genetics of the disease and how to manage the disease at home [17]. Educating parents on the genetics of sickle cell disease is essential as uncovered by this study. Parents were educated on what the disease was, its causes, how the disease is inherited and how family history plays a role in the inheritance of the disease. This study showed that as part of comprehensive follow-up care for newborns diagnosed with SCD at birth, parents need to be taken through several counselling and educational sessions or programmes with trained SCD nurses or staff trained in genetic counselling. This is to provide information about the genetic makeup of the disease, the way the disease is inherited through the genes of biological parents and how the history of the disease in the family may play a role in families having a better understanding of the disease makeup.

### Objective 3-Parent’s Adherence to Follow-up Care Procedures

The third research question sought to explore parents’ attitudes towards follow-up care procedures, i.e., adherence or non-adherence. One major theme emerged from this section, namely adherence to routine clinic care.

Routine clinic visitation, regular physical examinations and recording of clinical and laboratory information of patients with SCD are important [17]. Routine clinical procedures for parents at the Korle Bu Teaching Hospital included routine lab tests, daily medication and routine clinic visitation for physical examinations.

It was discovered from the findings of the study that parents adhered to these routine clinic requirements due to their desire to see their children healthy as well as from their own experiences with the disease. This is in contrast with a study by Ingerski et al [20] on clinic attendance of youth with SCD on Hydroxyurea where it was found that family size, financial capacity, incidence, and non-incidence of painful episodes were reasons why the youth were more likely to attend clinic regularly.

There were however some parents who could not adhere to these routine procedures due to socio-economic challenges as well as the COVID-19 pandemic as shown by findings from the study. The findings of COVID-19 are similar to a study by Kenney et al [21] into how COVID-91 affected ambulatory services and comprehensive care for people with SCD which revealed that COVID-91 induced stress and anxiety contributed significantly to an increase in patients’ no-shows and patients rescheduling. It is therefore important for hospital administration to plan better to be able to adjust to continue to provide care for newborns whose parents may be too anxious or apprehensive to attend a clinic during a pandemic.

Sickle cell disease is known to cause financial difficulties for families of people with the disease [22] and financial difficulties were one of the reasons why parents could not adhere to the routine clinic visitation and lab tests as they could not afford it.

### Limitations of the study

One limitation of the study is that this study did not explore the role of psychosocial factors in influencing parents’ acceptance of positive results and their attitudes toward routine follow-up care.

Another limitation of this study is that the opinions of the clinic staff and other people involved in the provision of comprehensive care were not included. This may have given a more thorough insight into the experiences of parents of newborns at the clinic, especially concerning the counselling and education offered to parents.

## Conclusions

This study reiterates the point that parents play an important role in providing comprehensive follow-up care for newborns who have tested positive for SCD. Parents’ reactions to disclosure of positive NBS results as well as their acceptance of the results were influenced by their prior knowledge and or experience of the disease. This calls for more community engagement and education to create awareness about the disease. There is also a need to strengthen measures which provide psychological and socio-economic support to families attending the KBTH clinic. There is also a need to promote a community-based, family-led model for SCD care through training of primary healthcare workers which could in turn reduce the geographical and financial barriers to health access.

## What is already known on this topic

1. Early identification of newborns with SCD and the provision of follow-up care help reduce under 5 morbidity and mortality.
2. Parents report diverse feelings when given positive SCD results for their newborns.

## What this study adds

1. Prior knowledge of sickle cell disease impacts parents’ reactions to the initial disclosure of positive SCD results of their newborns.
2. Parents play a vital role in providing comprehensive follow-up care for newborns diagnosed with SCD at birth.

## Competing interest

The authors declare no competing interests. No funding was obtained from a third party for the study. Authors 1, 3 and 4 however are staff of the Paediatric Sickle Cell Clinic and provide care to patients and families of children with sickle cell disease.

## Author’s contribution

Irene Kanyoke and Faustina Hayford Blankson conceived the investigation and participated in the design, data collection, interpretation and final drafting of the manuscript. Catherine Segbefia and Diana Dwuma-Badu helped with data collection and analysis and guided the overall write-up.

## Data Availability

All relevant data are within the manuscript and its Supporting Information files.

## Acknowledgements

We wish to thank all patients and guardians of the Paediatric Sickle Cell Clinic of Korle Bu Teaching Hospital. We also wish to thank the following staff members of the clinic: Albert Peprah, Irene Odoom Brown, Ellen Nkansah, Nawal Harith, Ayesha Musah, and Samuel Bainson for all their efforts in helping with data collection. I also want to thank Bamenla Goka, Jennifer Welbeck and Mercy Ampadu from the Child Health Department of Korle Bu Teaching Hospital for their assistance. Lastly, we extend our appreciation to Augustine Asubonteng, Sarah Asante Agyei and Mrs Mary Ethel Lamptey from the Sickle Cell Foundation of Ghana (SCFG) for their help with data collation.

## Supporting information

**S1 Table. Summary of socio-demographic information of participants**

**S2 Table. Codebook**

**Table 1.** Summary of socio-demographic information of participants

| Archival number | Marital status of participant (s) |
| --- | --- |
| KBTHPSCCII001 | married |
| KBTHPSCCII002 | married |
| KBTHPSCCII003 | Not married/in relationship with child's father |
| KBTHPSCCII004 | married |
| KBTHPSCCII005 | married |
| KBTHPSCCII006 | Married |
| KBTHPSCCII007 | married |
| KBTHPSCCII008 | married |
| KBTHPSCCII009 | married |
| KBTHPSCCII010 | married |
| KBTHPSCCII011 | Not married |
| KBTHPSCCII012 | married |
| KBTHPSCCII013 | Separated |
| KBTHPSCCII014 | Not married |
| KBTHPSCCII015 | married |
| KBTHPSCCII016 | married |
| KBTHPSCCII017 | married |
| KBTHPSCCII018 | married |
| KBTHPSCCII019 | Not married |
| KBTHPSCCII020 | separated |
| KBTHPSCCII021 | married |
| KBTHPSCCII022 | Not married. In relationship with baby father |
| KBTHPSCCII023 | Married |
| KBTHPSCCII024 | Married |

**Table 2.** Codebook

| Research Question/objective<br>(Open coding) | Themes<br>(Quotes from interviews) | List of Codes | EmergEd<br>Themes | Attitude<br>(From<br>Theoretical<br>framework) | Construct<br>(From SEM) |
| --- | --- | --- | --- | --- | --- |
| Reactions to the disclosure of positive results | *"You know, life is hard now, and I was scared that if it happens that she is a sickler, what would I do?"*<br>*"Because that first time she told me, I got scared. That's why I didn't believe it. I was worried".*<br>*"So him ((pointing to child with SCD at clinic)), when his results came, I wasn't surprised because I knew the dad is SC. so I wasn't surprised at all".*<br>*'he still doesn't believe. And I tell him that, if he didn't have it, the nurses wouldn't have told me he has it".*<br>*"It did help. Because, even before we got married, I was contemplating. And then when the news came, even though it came as a shock"* | 1. Scared<br>2. Worried<br>3. Not surprised<br>4. Disbelief<br>5. Surprised/ Shocked | 1. emotional response after disclosure | Influences | Intrapersonal (individual) |
| Reactions to the disclosure of positive results | *"well, I didn't want anyone to know. I just wanted it to be between me and my husband so I didn't tell anyone"*<br>*"nobody oo. I've not told anybody. Because of the stigma, I don't want to, to tell anybody"*<br>*"Yes. I told the caretaker. Because if I have to leave, and you don't give the medication, I will blame her. And then recently, I told my mother-in-law... and then my parents, my siblings are also aware".* | 1. Shared results with partner only<br>2. Fear of social exclusion stigma<br>3. Shared results with family and friends | 2. Decision to share results | Influences | Intrapersonal |
| Response to In-clinic counselling | *"How to how to, eh, is it, prevent it, eh, in terms of not to take something cold. And all those kinds of things. You will know the way when it's cold, you should put something, like cardigan, all those kinds of things. And you have to take your medication always, no crises, for the 0 months to 5 years. You won't have any problem".*<br>*"When they are babies, maybe you will see one of their hands swelling or their feet swollen. Or their eyes. and when they get temperature, you have to bring them to the hospital and not stay in the house. It might be infection".* | 1. Preventive measures/care at home (nutrition, hydration, warm clothes)<br>2. Awareness of treatment options<br>3. Awareness of possible complications<br>4. Danger signs | 3. Care and Management of Newborns with SCD | Network (with clinic staff) | Interpersonal |
| Response to In-clinic education | *"Yes. She said like, it was from both my husband and I. and that I have S in my blood and my husband also has S in his blood. And that's how come the child has the S".*<br>*"They took me through the sickling. How it forms. How their blood cells look like. Is it the white one or the, the red one? Yea, that looks like a bofloat, and theirs is like an arch, the moon, half moon, which is very hard not like the others that are soft that can pass through all the blood. Theirs because is hard, sometimes, it gets broken and chokes the veins that you have to give them the (massage) and all that. so, I was taught all that"*<br>*"It's me and our last born (meaning youngest sibling) who have the disease in our family".* | 1. Inheritance of SCD<br>2. Blood disorder<br>3. Family history of disease | 4. Knowledge of genetics of SCD | Network with clinic staff | Interpersonal |
| Attitudes towards Follow up procedures | *"I've been coming to clinic for the past 7 to 8 months. Every month. I'm here. Yes, sometimes even 2 weeks. In a month, I can come like 2 times".*<br>*"Because I have that experience. So I don't want her to pass through the stress I have gone through, so I come to clinic".*<br>*"We come. Every 3 months, we come. But ...we were asked to come and do some labs. But we couldn't do it. Because of the COVID. I was scared so I didn't come.*<br>*"Madam please, you see, for my son, I love him. And it's the doctor who knows, so whatever they tell me I have to listen and do it. But sometimes, when I'm unable to come on the date, it's usually because of financial difficulties. I may be lacking money for the labs .... But financially, it's been difficult. Because of money issues. I was supposed to bring him last month but because I didn't have money, I couldn't come".*<br>*"And so because of that, I don't play with the medication. Once it finishes, I make sure I refill it"*<br>*"oh it's usually the lab we frequently do"* | 1. Adherence to routine clinic visits (due to expectation of healthy outcome for child)<br>2. Adherence due to parents' personal experience with disease)<br>3. non-adherence to clinic visits due to COVID<br>4. non-adherence due to financial difficulty<br>5. Adherence to daily medication<br>6. routine lab tests | 5. Adherence to routine clinic | Influences | Intrapersonal |

## Notes

### Competing Interest Statement

The authors have declared no competing interest.

### Funding Statement

The author(s) received no specific funding for this work.

### Author Declarations

INSTITUTIONAL APPROVAL: KORLE BU TEACHING HOSPITAL-SCIENTIFIC AND TECHNICAL COMMITTEE/INSTITUTIONAL REVIEW BOARD (KBTHSTC/IRB/00094/2021)

## References

1. Asare EV, Wilson I, Benneh-Akwasi KAA, Dei-Adomakoh Y, Sey F, and Olayemi E. Burden of Sickle Cell Disease in Ghana: The Korle-Bu Experience. Advances in Hematology. 2018; 2018(1):1–5.

2. Russo G, De Franceschi L, Colombatti R, Rigano P, Perrotta S, Voi V et al. Current challenges in the management of patients with sickle cell disease - A report of the Italian experience. Orphanet Journal of Rare Disease. 2019; 14(1):1–9.

3. Hsu L, Nnodu EO, Brown BJ, Tluway F, King S, Dogara LG et al. White Paper: Pathways to Progress in Newborn Screening for Sickle Cell Disease in Sub-Saharan Africa. Journal of Tropical Disease. 2018; 06(02):1–10.

4. Archer NM, Inusa B, Makani J, Nkya S, Tshilolo L, Tubman VN et al. Enablers and barriers to newborn screening for sickle cell disease in Africa: Results from a qualitative study involving programmes in six countries. BMJ Open. 2022; 12(3):1–10.

5. Segbefia C, Goka B, Welbeck J, Oppong S, and Odame I. Implementing newborn screening for sickle cell disease in Accra, Ghana: Results and challenges. Pediatric Blood and Cancer. 2018; 66(S2):S266.

6. Osei-Akoto A, Lamptey M, Tetteh H, Ansong D, Paintsil V, Dennis-Antwi J et al. Newborn screening for sickle cell disease in Ghana -20 years of testing, tracking and followup. Vox Sanguinis. 2017; 112(S1):42.

7. Therrell BL, Lloyd-Puryear MA, Ohene-Frempong K, Ware RE, Padilla CD, Ambrose EE et al. Empowering newborn screening programs in African countries through establishment of an international collaborative effort. Journal of Community Genetics. 2020; 11(3):253–268.

8. Sims AM, Bonsu KO, Urbonya R, Farooq F, Tavernier F, Yamamoto M et al. Diagnosis patterns of sickle cell disease in Ghana: a secondary analysis. BMC Public Health. 2021; 21(1):1–7.

9. Segbefia C, Goka B, Welbeck J, Amegan-Aho K, Dwuma-Badu D, Rao S et al. Implementing newborn screening for sickle cell disease in Korle Bu Teaching Hospital, Accra: Results and lessons learned. Pediatric Blood Cancer. 2021; 68(7):1–7.

10. Dwuma-Badu D, Segbefia C, Kanyoke I, Peprah A, Odoom-Brown I, Harith N et al. Follow up of babies identified through newborn screening for sickle cell disease: Implementation of a nurse-led care pathway at a teaching hospital in Ghana. 18^th^ Annual Academy for Sickle Cell and Thalassaemia conference, London. 2023; 8–9. Available: https://www.ascatconferences.com/abstracts. Accessed on December 1, 2023.

11. Francis J, Johnston M, Robertson C, Glidewell L, Entwistle V, Eccles M et al. What is an adequate sample size? Operationalising data saturation for theory-based interview studies. Psychology and Health. 2010; 25(10):1229–1245.

12. Chudleigh J, Buckingham S, Dignan J, O’Driscoll S, Johnson K, Rees D et al. Parents’ Experiences of Receiving the Initial Positive Newborn Screening (NBS) Result for Cystic Fibrosis and Sickle Cell Disease. Journal of Genetic Counselling. 2016; 25(6):1215–1226.

13. Niekerk VK. Knowledge and experiences of parents with children affected by Sickle Cell Disease in Cape Town. University of Cape Town. 2015. Available: https://open.uct.ac.za/items/95366deb-2ff5-49db-8a61-583cb204da5e. Accessed on May 31, 2024.

14. Ulph F, Cullinan T, Qureshi N, and J. Kai. Parents’ responses to receiving sickle cell or cystic fibrosis carrier results for their child following newborn screening. European Journal of Human Genetics. 2015; 23(4):459–465.

15. Adegboyega LO. Counselling Needs of Sickle-Cell Anaemia Adolescents in Ekiti State, Nigeria. Ethiopian Journal of Health Sciences. 2020; 30(6):1005–1010.

16. Makani J, Ofori-Acquah SF, Nnodu O, Wonkam A and Ohene-Frempong K. Sickle cell disease: New opportunities and challenges in Africa. The Scientific World Journal 2013; 2013: 1–16.

17. Johnso PL, Lombardo FA, and Moore CD. Options in the management of sickle cell disease. U.S. Pharmacist. 2015; 40(7):4–7.

18. Colah RB, Mehta P, and Mukherjee MB. Newborn screening for sickle cell disease: Indian experience. International Journal of Neonatal Screening. 2018; 4(4):1–8.

19. King L, Fraser R, Forbes M, Grindley M, Ali S, and Reid M. Newborn sickle cell disease screening: The Jamaican experience (1995-2006). Journal of Medical Screening. 2007; 14(3):117–122.

20. Ingerski LM, Arnold TL, Banks G, Porter JS, and Wang WC. Clinic Attendance of Youth with Sickle Cell Disease on Hydroxyurea Treatment. Journal of Pediatric Hematology/Oncology. 2017; 39(5): 345–349.

21. Kenney MO, Becerra B, Beatty SA, and Smith W. Physicians’ opinions of covid-19 ambulatory care constraints: A survey of sickle cell clinicians. Journal of Ambulatory Care Management. 2012; 44(4):325–330.

22. da Silva De Jesus AC, Konstantyner T, Lôbo IKV, and Braga JAP. Socioeconomic and nutritional characteristics of children and adolescents with sickle cell anemia: A systematic review. Revista Paulista de Pediatria. 2018; 36(4):491–499.

